# *APOE*\*4 Risk-Modifying Genes and Drug Targets in Alzheimer’s Disease through Cell-Type Specific Genomic Analyses

**DOI:** 10.64898/2025.12.02.25341367

**Authors:** Youjie Zeng, Noah Cook, Chenyu Yang, Sathesh K. Sivasankaran, Masashi Fujita, Zachary A. Gardell, Yann Le Guen, Daichi Shigemizu, Kouichi Ozaki, Takashi Morizono, Norikazu Hara, Akinori Miyashita, Takeshi Ikeuchi, Cyril Pottier, Carlos Cruchaga, Valerio Napolioni, M. Ryan Corces, Vilas Menon, Michael D. Greicius, Michael E. Belloy

## Abstract

**Importance:** *APOE*\*4 is the strongest, common genetic risk factor for late-onset Alzheimer’s disease (AD) with widespread and cell-type-specific impact on disease pathogenesis. Genetics and omics studies can help identify genes that counteract the effects of *APOE*\*4, but so far have remained relatively small and crucially did not assess genetic findings through a cell-type-specific framework.

**Objectives:** Perform a large-scale *APOE*\*4 stratified genome-wide association study (GWAS) of AD integrated with genetically tethered cell-type-specific multi-omics data.

**Design:** Meta-analysis of *APOE*\*4 stratified AD GWAS in case-control, family-based, population-based, and longitudinal AD-related cohorts from the Alzheimer’s Disease Genetics Consortium, Alzheimer’s Disease Sequencing Project, and UK Biobank. Integration of GWAS with brain cell-type-specific genetic regulation of gene expression data, from the Religious Orders Study and Memory and Aging Project, to identify *APOE*\*4 and cell-type-specific AD genes. Cell-type-specific multi-omics gene prioritization followed by compound and drug repurposing. Data analyzed between January 2023 and September 2025.

**Setting:** Genetic data available from high-density single-nucleotide polymorphism (SNP) microarrays and whole-genome sequencing (WGS). Single-nucleus (sn) RNA-seq data from dorsolateral prefrontal cortex.

**Participants:** 567,521 eligible participants for AD genetic association studies were selected from referred and volunteer samples, of which 119,852 were excluded for analysis exclusion criteria.

**Main Outcome and Measures:** *APOE*\*4 and cell-type-specific gene Z-scores and FDR-corrected P-values. Gene prioritization scores and *APOE*\*4 stratified enriched compounds.

**Results:** 67 and 17 significant cell-type-gene pairs were identified in *APOE*\*4 non-carriers (*APOE*\*4-) and carriers (*APOE*\*4+) respectively. Oligodendrocytes displayed the largest proportion of *APOE*\*4+ genes. 46 cell-type-gene pairs were supported by at least half of the gene prioritization analyses. Several prioritized genes were druggable and displayed enrichment of *APOE*\*4 stratified drugs or compounds. Top *APOE*\*4+ genes with connections to enriched drugs or compounds included TNS3 (astrocytes), CISD1 and SLC23A2 (oligodendrocytes), and UBXN4 (excitatory neurons).

**Conclusion and Relevance:** We identified a set of *APOE*\*4 stratified genes that may be causal for AD through brain cell-type-specific mechanisms and prioritized top genes for further interrogation. We additionally identified compounds that may be repurposed or shed light on therapeutic avenues for treating AD based on an individual’s *APOE*\*4 status. Top identified compounds such as Hydrocortisone and Trolox implicate oxidative stress and neuroinflammation as potential biological targets in *APOE*\*4+ individuals.

## Introduction

The *Apolipoprotein* E ε4 allele (*APOE*\*4) is the strongest, common genetic risk factor for late-onset Alzheimer’s disease (AD), with one and two copies respectively increasing risk about 4 and 12-fold in European-ancestry populations^1^. While the effect magnitude of *APOE*\*4 varies across ancestries, it consistently represents the most prominent genetic risk factor for AD^2^. Despite consensus that countering *APOE*\*4 represents an important therapeutic objective for AD, few drugs or clinical trials directly target *APOE*\*4^3–6^. The challenge in part stems from the highly pleiotropic nature of *APOE*\*4 and uncertainty about where and how to target it^1^. Importantly, recent work highlights that the impact of *APOE*\*4 on AD pathobiology differs across brain cell-types, suggesting that cell-type informed approaches may be crucial to identify genes that can counter the effect of *APOE*\*4^7–14^.

Genome-wide association studies (GWAS) have been instrumental in identifying AD risk variants and genes^15,16^, paving the way for novel therapeutic targets^17^. Few prior GWAS and targeted genetic analyses further considered *APOE*\*4 status, uncovering promising genes such as *Klotho* and *Fibronectin1* that may counter *APOE*\*4^18–21^, but novel, larger *APOE*\*4 stratified AD GWAS are urgently needed. Historically, bulk brain tissue transcriptomics datasets have enabled linking genetic variants to gene expression (expression quantitative trait loci, eQTLs), which, in turn, have supported mapping genetic signals from AD GWAS to their likely causally related genes^15,22,23^. Recent advances in single-cell and single-nucleus RNA sequencing (snRNA-seq) enable studying the genetic regulation of gene expression at cell-type resolution (sc-eQTL), revealing many regulatory signals that were missed in bulk eQTL analyses^24–26^.

To address these gaps, we conducted a meta-analysis of *APOE*\*4 stratified AD GWAS across multiple European-ancestry cohorts and integrated findings with a large brain sc-eQTL resource to conduct an *APOE*\*4 stratified cell-type-specific transcriptome-wide association study (cTWAS). This approach identifies genetic signals for AD that both depend on *APOE*\*4 status and regulate gene expression in specific brain cell-types. Using additional multi-ancestry GWAS samples and various cell-specific multi-omics follow-up analyses, we further prioritized the most promising genes and leveraged them to identify potential *APOE*\*4 stratified drug targets.

## Methods

The current study followed STREGA reporting guidelines. Participants or their caregivers provided written informed consent in the original studies. The study protocol was granted an exemption by the Washington University Institutional Review Board because the analyses were carried out on “de-identified, off-the-shelf” data; therefore, additional informed consent was not required.

### Genetic Data Ascertainment & Processing

Case-control, family-based, and population AD genetic cohorts from the Alzheimer’s Disease Genetics Consortium (ADGC) and Alzheimer’s Disease Sequencing Project (ADSP) were available from public repositories and contributed GWAS data for European (EUR) and African admixed (AFR) ancestry subjects (**eFigure-1**) through whole genome sequencing (WGS) or imputed SNP microarrays (**eTable-1-2**)^27,28^; subjects were clinically diagnosed with 40% of cases pathology confirmed (**eTable-3**). UK Biobank (UKB) contributed EUR AD GWAS data via imputed SNP microarrays^29^; AD phenotypes were determined from health-registry records or questionnaire-identified proxy-cases whose parents were affected by AD or dementia^15,30^. Japanese AD GWAS data were available through imputed SNP microarrays from the National Center for Geriatrics and Gerontology and Niigata University (**eTable-4**)^31^; subjects were clinically diagnosed. Details on GWAS processing and variant metrics are in the **eMethods** and **eTables-5-7**.

### Genetic Statistical Analyses - *APOE\**4 stratified GWAS and cTWAS

GWAS were performed separately in *APOE*\*4 non-carriers (*APOE*\*4-) and carriers (*APOE*\*4+), evaluating AD logistic regressions, adjusted for sex, age, *APOE*\*2 and *APOE*\*4 dosage, technical covariates, and genetic principal components, as applicable per dataset. EUR samples accounted for the majority of data, with GWAS conducted using mixed models (BOLT-LMM v2.4)^32^, followed by fixed-effects meta-analyses (GWAMA)^33^. Variants were intersected with those present in ADGC and a genotyping rate ≥90% across cohorts to increase AD specificity^34,35^. Additional *APOE*\*4 stratified GWAS were conducted in AFR and Japanese cohorts (Plink v2.0)^36^. Effective sample sizes were calculated using N_Eff_ =4*v*(1-v)^37^, where v=N_Cases_ /(N_Cases_ +N_Controls_), with contributions from UKB further divided by 4 to account for proxy-GWAS^30^.

*APOE*\*4 stratified cTWAS were performed by integrating EUR *APOE*\*4 stratified GWAS findings with snRNA-seq data, paired with WGS data, from the dorsolateral prefrontal cortex of 424 EUR participants in the Religious Orders Study (ROS) and Memory and Aging Project (MAP). Data processing is described in Fujita et al.^24^ Briefly, seven major cell-types were available to link genetic variants to gene expression. For each cell-type, sc-eQTL analyses were conducted (Matrix eQTL v.2.3)^38^ and transcriptogenomic weights were generated through FUSION for genes that displayed significant heritability (P<0.05) in their cis windows (± 1Mb around transcription start site)^39^. The resultant cell-type-specific variant-gene weights were used to conduct *APOE*\*4 stratified cTWAS and identify cell-type-gene pairs likely causal to AD (cf. **eTable-8** for gene counts per cell-type). To ensure findings were specific to a given *APOE*\*4 stratum, genes were filtered to a false discovery rate P_FDR_<0.05 in one stratum and either (1) a non-significant association (P>0.05) with the same effect direction in the other stratum, or (2) an opposite effect direction in the other stratum. Additionally, for loci with multiple significant genes, if the most significant gene was not biased to a given *APOE*\*4 stratum, as in the above criteria, all genes at the locus were excluded.

### Gene Prioritization

*APOE*\*4 heterogeneous cTWAS candidate genes were further assessed using a series of multi-omics approaches to validate findings within their corresponding cell-type and *APOE*\*4 stratum. Increasing levels of support were used to prioritize the most promising genes.

#### Multi-ancestry cTWAS

To assess cross-ancestry concordance of *APOE*\*4 heterogeneity, EUR cTWAS genes were evaluated through equivalent FUSION analyses with respective AFR and Japanese *APOE*\*4 stratified GWAS findings. Meta-analyses were conducted through sample-size weighted combination of Z-scores. Cell-type-gene pairs were considered consistent if the P-value in the respective *APOE*\*4 stratum (e.g. *APOE*\*4+) improved in cross-ancestry meta-analysis, while the opposite *APOE*\*4 stratum (e.g. *APOE*\*4-) maintained P>0.05. Few genes could be assessed in the Japanese cTWAS, likely due to limited variant and linkage disequilibrium (LD) overlap with EUR data, so these results were not used for gene prioritization.

#### Colocalization analysis

Consistent with TWAS and cTWAS follow-up approaches in prior studies^40–43^, genetic colocalization (COLOC) analyses were conducted for cell-type-gene pairs of interest across the AD GWAS and ROSMAP sc-eQTL data. The goal of these analyses is to evaluate support for a causally shared variant (posterior probability PP4) which helps corroborate a causal link for the respective gene to AD. COLOC was performed using (1) standard approximate Bayes factor (ABF) analyses^44^, (2) ABF analyses with priors adjusted to reflect increased probabilities of variants being associated with both AD and gene expression (i.e. the conditions for cTWAS discoveries), and (3) SuSiE-based COLOC^45^, which accounts for scenarios where multiple causal variants are present at the locus. Across analyses, the best PP4 value was used with values ≥0.4 and ≥0.7 respectively marking suggestive and strong COLOC support.

#### Summary-data-based Mendelian randomization (SMR)

Consistent with TWAS and cTWAS follow-up approaches in prior studies^40–43^, SMR analyses were conducted for cell-type-gene pairs of interest across the AD GWAS and ROSMAP sc-eQTL data. The goal of SMR is to assess the causal effects of gene expression on AD using the top cis sc-eQTL variant as an instrumental variable, while the HEIDI test assesses if the association is driven by LD^46^. Cell-type-gene pairs with P_FDR_ <0.05 and P_HEIDI_ >0.05 were considered to have SMR support.

#### scATAC-seq validation

For all cell-type-gene pairs supported by COLOC, the top colocalizing variants and those in close LD (R^2^>0.8) were assessed for overlap with regulatory peaks from single-cell ATAC-seq data in matching cell-types from human brain tissues^47^. These analyses aim to corroborate that the cTWAS-identified cell-type-specific genetic signals may manifest via cell-type-specific epigenetic mechanisms.

#### Single-cell Differential Expression of Genes (scDEG)

Pseudobulk expression matrices for seven brain cell-types from ROSMAP snRNA-seq data were count-normalized using the trimmed mean of M-values (TMM) in edgeR^24,48^. scDEG analyses were performed with limma^49^, adjusting for sex, post-mortem interval, age-at-death, APOE*2 and *APOE*\*4 dosage, and batch. Outcome phenotypes comprised both binary and global quantitative metrics of clinical and pathological AD (cf. **eMethods**). Analyses were conducted in both non-stratified and *APOE*\*4 stratified settings. In non-stratified scDEG, support was defined as P<0.05 in the respective cell-type-gene pair. In *APOE*\*4 stratified scDEG, support required (i) P<0.05 in the respective *APOE*\*4 stratum and cell-type-gene pair and (ii) an opposite effect direction or absolute log-fold change ≥1.5 larger compared to the opposite stratum. These analyses aim to corroborate a direct gene association with AD dementia or pathology within the respective cell-types and *APOE*\*4 strata.

### Drug Target Analyses

*APOE*\*4 stratified cTWAS genes prioritized through more than half of the follow-up analyses were intersected with a list of 4,479 druggable genes from Finan et al. to identify Tier-1 druggable cTWAS genes^50^. Remaining prioritized cTWAS genes were expanded into networks of AD-related genes through EpiGraphDB^51^, with expanded genes restricted to those with evidence of druggability^50^. *APOE*\*4 stratified compound and drug enrichment analysis were performed on the expanded gene lists using the DSigDB drug signature database (clusterProfiler package)^52,53^. Compounds and drugs were considered significantly enriched at P_FDR_<0.05 and further filtered to those targeting at least 2 prioritized genes to increase robustness.

## Results

The study design is summarized in **Figure-1**. *APOE*\*4 stratified GWAS included 432,508 EUR, 7,270 AFR, and 7,891 Japanese individuals (**eTables-3-4**) and revealed a novel *APOE*\*4+ (*RBM6*) and *APOE*\*4-locus (*TECPR2*) in EUR individuals (**eFigure-2**; all loci **eTable-7**). There were no signs of genomic inflation (**eFigure-3**). By integrating our GWAS with genetically tethered brain snRNA-seq data to perform *APOE*\*4 stratified cTWAS, we identified 84 cell-type-gene pairs that additionally passed *APOE*\*4 heterogeneity criteria (**Table-1**; **Figure-2**; **eFigure-4**; full results **eTables-8-10**). These involved 59 unique genes–45 *APOE*\*4- and 14 *APOE*\*4+ genes–across 42 genomic loci of which 29 were novel compared to prior AD GWAS. The MAPT locus, known for its broad and complex LD structure^54^, contained the highest number of *APOE*\*4-candidate genes across multiple cell-types. Most genes (n=45) were however identified in only one cell-type (**Figure-3A**). Oligodendrocytes exhibited the highest proportion of *APOE*\*4+ genes (**Figure-3B**; **eFigure-5**) of which three genes were respectively related to vesicular transport, mitochondrial metabolism, autophagy, and iron homeostasis pathways (**eTable-11;** cf. **eMethods**). Locus plots for cell-type-gene pairs showcase *APOE*\*4 heterogeneity of regional AD genetic signals (**eFigure-6**). Top AD variants across loci displayed strong correlation of effect sizes across ADGC/ADSP and UKB (ρ=0.90, R^2^=0.81), confirming findings were not biased by using proxy-GWAS in UKB (**eFigure-7**)^35,55^.

**Table 1.**
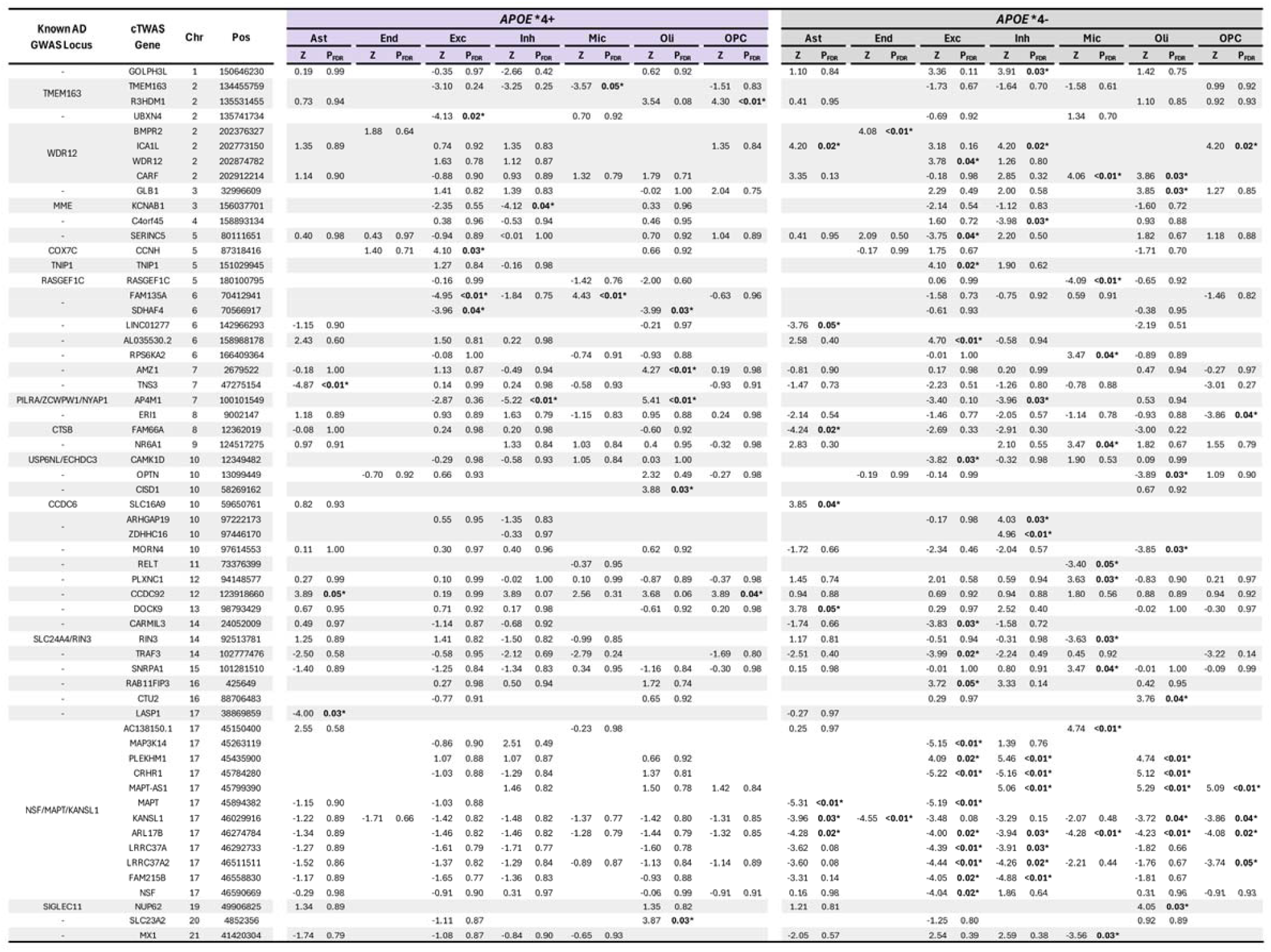
Significant Alzheimer’s disease associated cell-type-gene pairs exhibiting *APOE*\*4 specificity. Positive Z-scores indicate increased gene expression increases AD risk while negative Z-scores scores indicate increased gene expression decreases AD risk.

**Figure 1.**
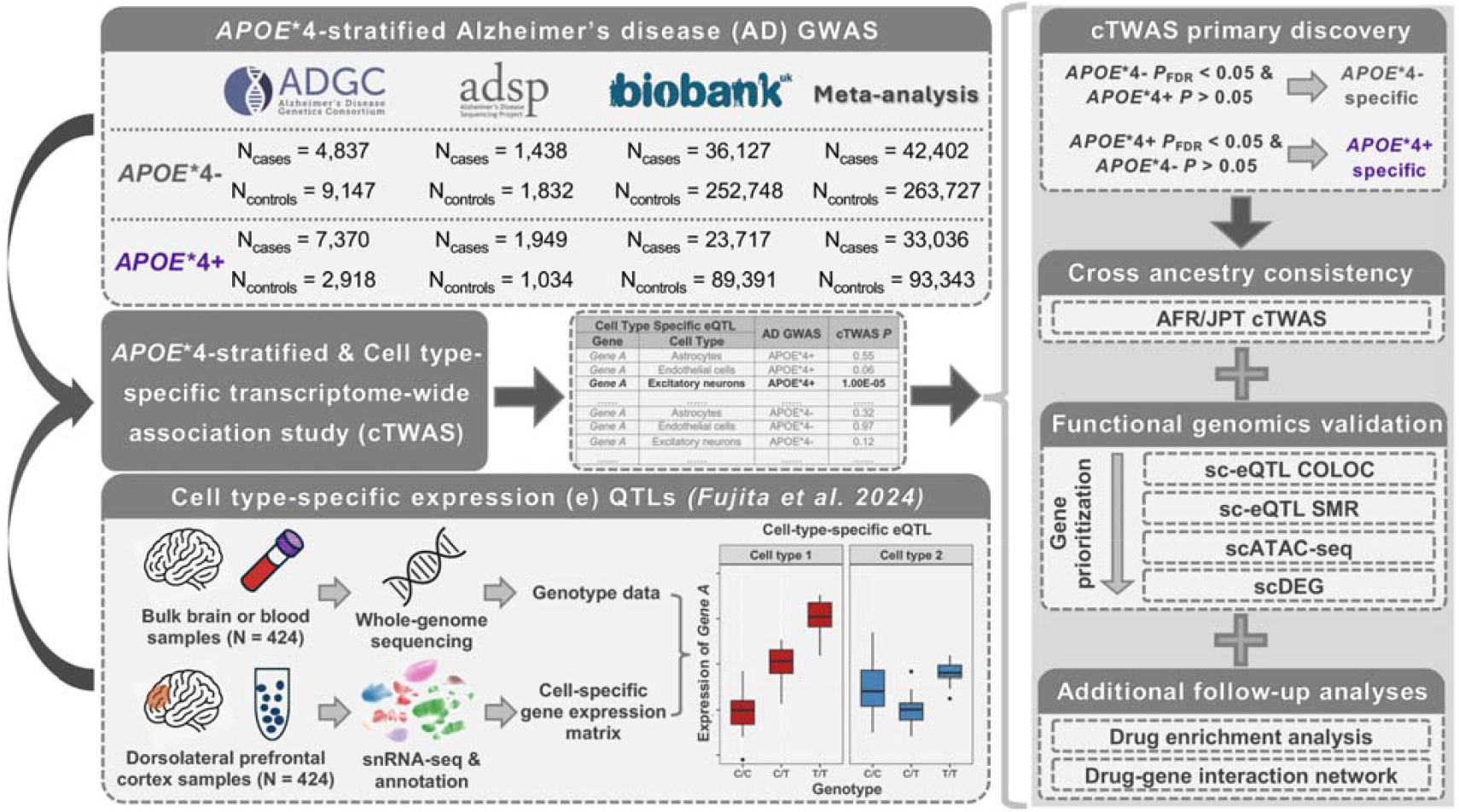
Schematic overview of the study design. **Left)** *APOE*\*4 stratified GWAS of AD in European ancestry subjects were integrated with snRNA-seq and paired whole genome sequencing data from ROSMAP brain tissue to perform an *APOE*\*4 stratified cTWAS. This approach implicates cell-type-gene pairs that are likely causal to AD in an *APOE*\*4 dependent manner. Right top) cTWAS findings were filtered for *APOE*\*4 specificity by retaining only significant (P_FDR_<0.05) genes in a given *APOE*\*4 stratum that were not significant (P>0.05) or showed discordant effect directions in the opposite stratum. Right middle) Following cTWAS gene discoveries, multiple types of gene prioritization analyses were conducted, including cross-ancestry cTWAS, leveraging smaller AD genetic datasets in AFR and Japanese cohorts to assess consistent APOE*4 heterogeneity effects, and a series of cell-type-specific multi-omics analyses. Right bottom) Top prioritized genes were used to perform drug repurposing analyses. Abbreviations: Alzheimer’s Disease Genetics Consortium, ADGC; Alzheimer’s disease Sequencing Project, ADSP; UK Biobank, UKB; Religious Orders Study and Memory and Aging Project, ROSMAP; genome-wide association study, GWAS; cell-type-specific transcriptome-wide association study, cTWAS; African, AFR; Japanese in Tokyo, JPT; single-cell expression quantitative trait locus, sc-eQTL; genetic colocalization analysis, COLOC; summary-based mendelian randomization, SMR; single-cell assay for transposase-accessible chromatin using sequencing, scATAC-seq; cell-type specific differential expression of genes, scDEG.

**Figure 2.**
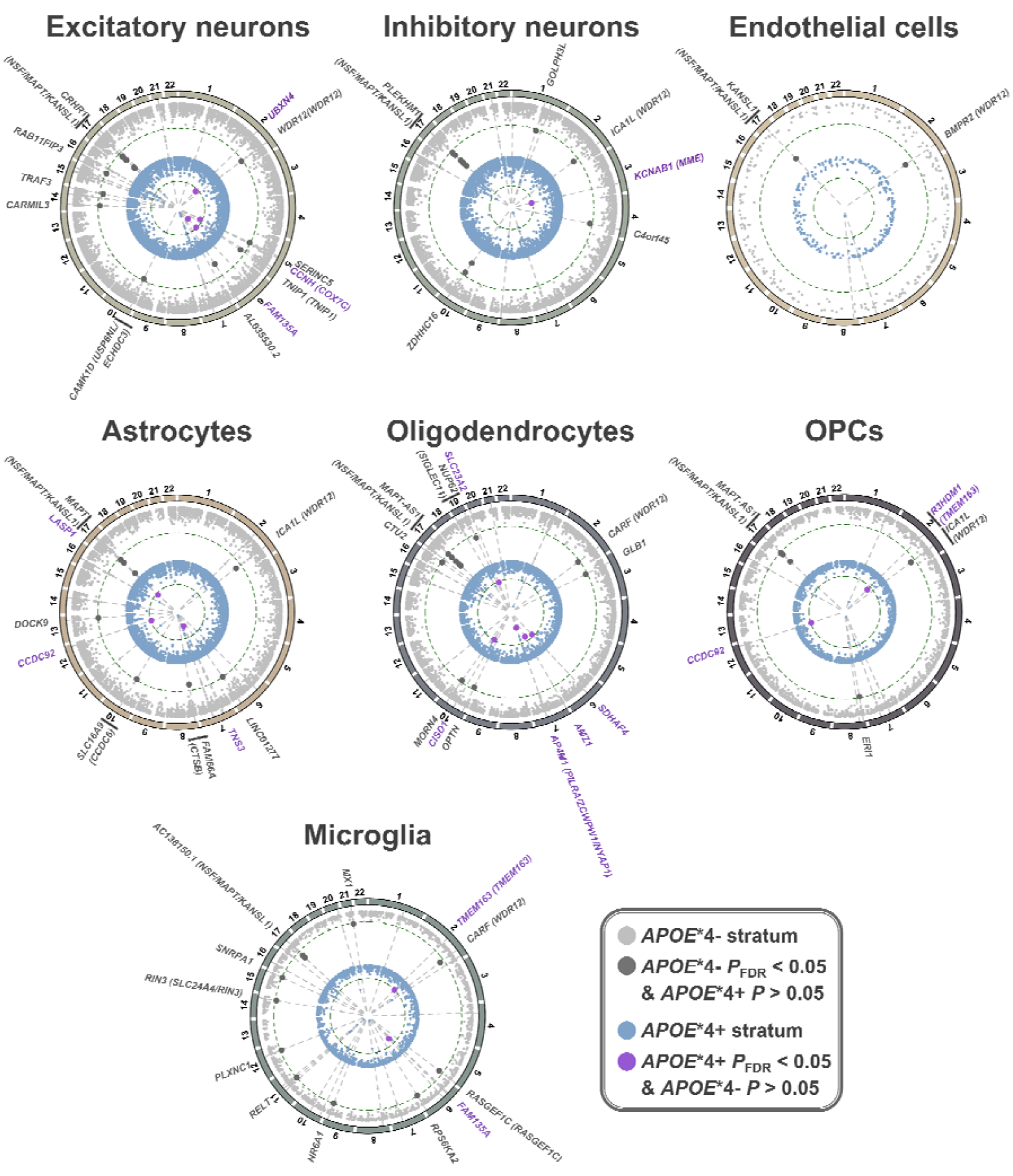
*APOE*\*4 stratified cell-type-specific transcriptome-wide association study (cTWAS) findings. For each cell-type, a circos plot is shown, depicting *APOE*\*4-findings on the outer layer and *APOE*\*4+ findings on the inner layer. The radial axes indicate the -log_10_(P-value), i.e. significance of gene association, with increasing significance toward the center of the plots. Dashed circular lines indicate P_FDR_<0.05 thresholds. Significant associations that displayed evidence of *APOE*\*4 specificity (dotted lines show alignment across *APOE*\*4 strata) are labeled with unique colors (cf. legend) and their respective gene names, including further gene annotation if they fell within known AD risk loci (genes between parentheses). If multiple genes were located within the same locus (within a 1 Mb window), only the top significant gene is labeled in the outer track.

**Figure 3.**
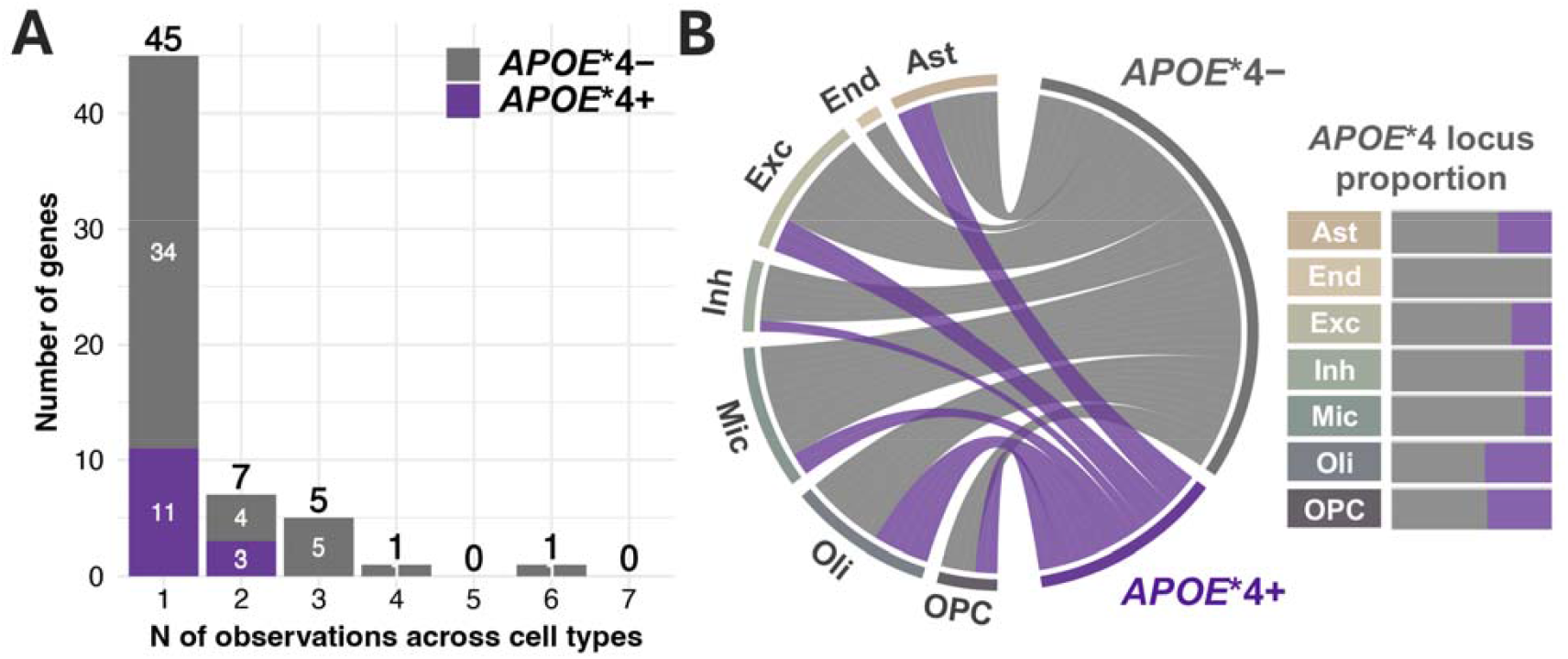
Cell-type-specificity of *APOE*\*4 stratified cTWAS results. **A)** The distribution of respective gene observations across cell-types is shown, indicating that most genes were observed in only one cell-type. **B)** Chord diagram (left) illustrates AD risk loci across *APOE*\*4 and cell-type groups, while the matching distributions are visualized in bar graphs (right). Notably, Oligodendrocytes showed the highest proportion of *APOE*\*4+ risk modifying loci.

Candidate genes from cTWAS were subjected to 6 gene prioritization analyses (**eTables-12-18**; **eFigure-8**) summarized in **Figure-4A** and **eTable-19** (findings excluding MAPT locus: **eFigure-9, eTable-20**). Overall, 11/17 *APOE*\*4+ (**Figure-4B**) and 35/67 *APOE*\*4-cell-type-gene pairs (**eFigure-10A**) were supported by more than half of the prioritization analyses. The druggability of these prioritized genes and their connected AD-related genes is presented in **eTable-21**. Using these gene sets, we identified 106 and 786 enriched compounds or drugs in *APOE*\*4+ and *APOE*\*4-strata, respectively (**eTable-22**). In extended analyses excluding the *APOE*\*4-prioritized gene TRAF3, which had extensive AD-gene-drug interactions, 101 compounds remained significantly enriched in *APOE*\*4-analyses (**eTable-22**). The top 10 *APOE*\*4+ compounds and their gene interactions highlighted promising drug targets including *TNS3, CISD1, SLC23A2*, and *UBXN4* (**Figure-4C-D**; *APOE*\*4-findings: **eFigure-10B-C**). Higher TNS3 expression in astrocytes was protective of AD in *APOE*\*4+ subjects and its lead genetic signal fell within a distinct astrocyte snATAC-seq peak (**eFigure-11**). Higher *CISD1* expression in oligodendrocytes increased AD risk in *APOE*\*4+ subjects. Leveraging independent plasma proteomics data, we additionally replicated the observation that CISD1 levels were significantly increased in *APOE*\*4+ (but not *APOE*\*4-) AD cases versus controls (**eTable-23**; cf. **eMethods**).

**Figure 4.**
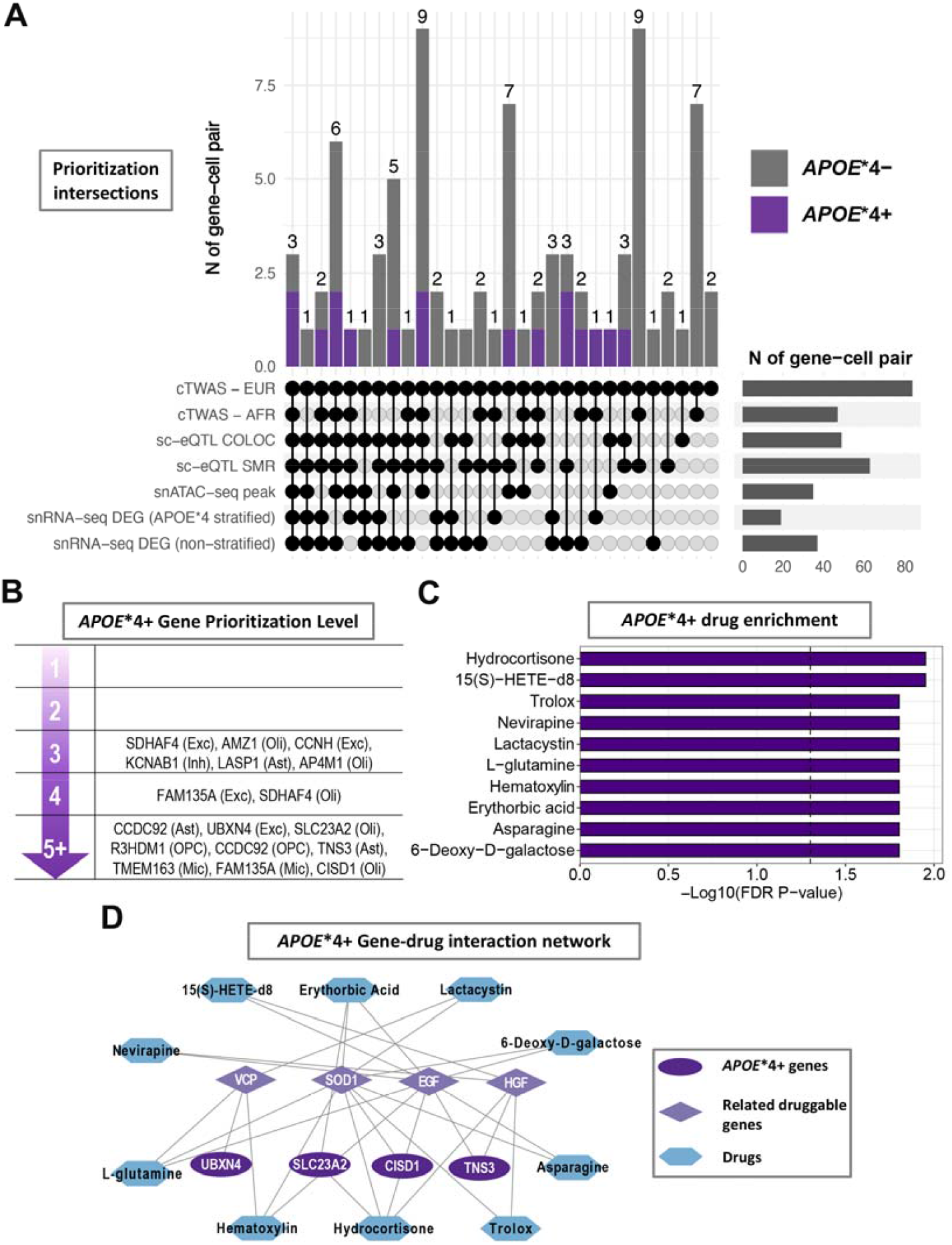
*APOE*4 s*tratified gene and drug target prioritization. **A)** The UpSet plot summarizes the European cTWAS significant cell-type-gene pairs with their varying supporting gene prioritization findings, arranged highest to lowest supporting observations from left to right. **B)** The schematic indicates cell-type-gene pairs with increasing levels of gene prioritization support in the *APOE*\*4+ stratum. **C)** Compound and drug enrichment results for *APOE*\*4+ specific genes (with prioritization level ≥ 4) and their interacting AD-related, druggable genes. The bar graph shows the top 10 FDR significant (vertical dotted line) compounds and drugs. D) Gene-drug interaction network matching to (C), including *APOE*\*4+ cTWAS genes and their interacting AD-related genes that were connected to the top 10 enriched compounds or drugs.

## Discussion

Our *APOE*\*4 stratified GWAS identified many loci previously associated with AD in non-stratified AD GWAS (**eFigure-2**)^15,16^, confirming the robustness of our genetic findings. Compared to prior *APOE*\*4 stratified AD GWAS by Jun et al. and Madrid et al.^56,57^, in which effective sample sizes ranged 18,130-23,412 in respective *APOE*\*4 strata, our GWAS was better powered with effective sample sizes of 38,608 and 50,752 in *APOE*\*4+ and *APOE*\*4-subjects, respectively. By leveraging our novel GWAS results to perform *APOE*\*4 stratified cTWAS, we further identified 13 known and 29 novel AD risk loci whose associations depended on *APOE*\*4 status and for which we could nominate potential causal cell-type-gene pairs. Considering recent reports that proxy-AD GWAS can confer biases to genetic association signals^58–60^, the *APOE*\*4 proxy-AD GWAS in UKB was performed according to recommendations from Wu et al. to reduce bias (cf. **eMethods**)^60^. Importantly, we further observed that effect sizes of AD genetic signals underlying the cTWAS findings were strongly correlated across ADGC/ADSP and UKB analyses, confirming gene discoveries were not driven by the use of proxy-AD GWAS. While we observed a larger number of *APOE*\*4-cTWAS loci (31 compared to 11 *APOE*\*4+), this appeared to be driven by the larger power in the *APOE*\*4-analyses (**eFigure-12**). Excitatory neurons, microglia, and oligodendrocytes were the most implicated cell-types by cTWAS, each respectively linked to 12 AD genetic loci displaying *APOE*\*4 heterogeneity. However, microglia had the fewest heritable genes–a prerequisite for gene inclusion in cTWAS–compared to excitatory neurons or oligodendrocytes (∼40%), suggesting a relative enrichment of microglial genes (**eTables-8-9**). This is consistent with prior reports that many AD genes are expressed by microglia^25,26,61,62^. Interestingly, oligodendrocytes displayed the largest proportion of *APOE*\*4+ genes, which remained robust even when applying stricter gene prioritization filters (**eFigures-13-14**). The relevance of oligodendrocytes in AD pathogenesis is increasingly recognized^63^, with recent work showing that *APOE*\*4 impairs myelination by dysregulating cholesterol metabolism in oligodendrocytes^11^.

Similar to Jun et al.^57^, we identified the *MAPT* locus in *APOE*\*4-subjects, for which their gene prioritization efforts nominated *KANSL1* rather than *MAPT*. In contrast, we identified 13 candidate genes (including *KANSL1*) and observed the strongest effect sizes for *MAPT* (astrocytes, excitatory neurons) and its anti-sense RNA, *MAPT-AS1* (inhibitory neurons, oligodendrocytes, oligodendrocyte precursor cells). Four of these cell-type-gene pairs were confirmed by more than half (≥4) of gene prioritization analyses, with the excitatory neuron-*MAPT* pair supported by only 3. While colocalization PP4 results for *MAPT/MAPT-AS1* were only suggestive, visual inspection confirmed strong colocalization (cf. **eFigure-6**), which reflects challenges conferred by the complex genetic LD structure at the *MAPT* locus^54^. Surprisingly, the cTWAS effect directions indicated that increased *MAPT* (and decreased *MAPT-AS1*) levels are protective of AD. Fujita *et al*. previously reported that most astrocyte and excitatory neuronal sc-eQTL findings in the ROSMAP cohort–used in our cTWAS–had concordant effect directions with those observed in iPSC-derived astrocytes and neurons, except a few notable ones, including *MAPT*^24^. To confirm the robustness of this observation, we queried findings from the recent SingleBrain resource^64^, which reprocessed data from *Fujita et al*. and three other snRNA-seq datasets–doubling sample sizes for sc-eQTL analyses–and observed replications across all datasets for all cell-type-gene pairs except *MAPT* in excitatory neurons (**eTable-24**). We further verified that the top (protective) *APOE*\*4-AD and (expression increasing) *MAPT* sc-eQTL signals fell on the large H2 inversion haplotype that has historically been associated with protection against AD^54^ (**eTable-25**). Altogether, we provide evidence that genetic regulation of *MAPT* expression across different brain cell-types has a stronger impact on AD risk in *APOE*\*4-versus *APOE*\*4+ subjects but surprisingly reveal a protective role for increased *MAPT* expression. While the origin of these observations remains unclear, they will represent an important topic of investigation for future studies. It should still be considered that multiple genes at the MAPT locus may be causal to AD^54^. Notably, *KANSL1* remains a strong candidate as it is the KAT8 Regulatory NSL Complex Subunit 1 that regulates KAT8, which has been implicated by multiple prior AD GWAS^15^.

We further focused on cTWAS genes with strong prioritization support and assessed which of these constitute promising drug targets. In *APOE*\*4-subjects, *TRAF3* was identified in excitatory neurons and its underlying genetic signal represented an independent association from the novel genome-wide significant *APOE*\*4-GWAS signal at *TECPR2* within the same locus (**eFigure-6.24**). *TRAF3* displayed extensive connections with known, druggable AD genes and, while *TRAF3* is primarily tied to immune cell function, it has also been implicated in mediating neuronal apoptosis^65^, corroborating its potential as a drug target for AD. Top *APOE*\*4+ drug targets included *TNS3* (astrocytes), *CISD1* and *SLC23A2* (oligodendrocytes), and *UBXN4* (excitatory neurons). *UBXN4* may impact AD through its role in endoplasmic reticulum protein degradation, while *SLC23A2* (a.k.a. *SVCT2*) is a Vitamin C co-transporter that may affect AD through its role in oligodendrocyte myelination^66^. A recent study implicated *TNS3* in microglial gene networks in glioblastoma and AD and reported an inverse correlation between *TNS3* and *APOE* expression levels^67^. This complements our observation of a protective effect of increased astrocyte *TNS3* levels in *APOE*\*4+ subjects and may corroborate the emerging consensus to reduce *APOE* levels in *APOE*\*4+ subjects to counter AD risk^3^. Further, TNS3 interacts with AD-related druggable genes EGF and HGF, which in turn interact with EGFR (**eFigure-11**), a gene that was recently implicated in Alzheimer’s disease through GWAS^15^ and itself shows strong potential as a dual drug target for cancer and AD^68^. For *CISD1*, we replicated its *APOE*\*4+ specific AD risk-increasing effect from cTWAS in an independent plasma proteomics dataset in which higher CISD1 levels were associated with AD risk solely in *APOE*\*4+ subjects. *CISD1* encodes an outer mitochondrial membrane protein that plays an important role in regulating oxidation^69^. Additionally, *CISD1*, as well as its connected druggable gene *SOD1*, are linked to ferroptosis which is increasingly implicated in microglial AD pathogenesis^61,70,71^. Importantly, SOD1 is a hallmark gene in amyotrophic lateral sclerosis (ALS) and while there is no direct link between AD and ALS, our findings suggest that converging mechanisms such as oxidative stress and protein misfolding may pinpoint drug repurposing opportunities for both diseases. Finally, we identified compounds or drugs that were significantly enriched in the prioritized gene sets. Among top compounds, Hydrocortisone and Trolox implicated oxidative stress and inflammation as biological targets that may confer increased benefit in *APOE*\*4+ versus *APOE*\*4-subjects. Considering that elevated endogenous cortisol has been linked to increased AD risk^72^, the enrichment of Hydrocortisone likely reflects a gene signature that aligns with reducing cortisol as a therapeutic target. Similarly, Trolox, an analog of Vitamin E, may protect against AD through reducing oxidative stress^73^.

## Limitations

Despite the large size of our GWAS, the lower power in *APOE*\*4+ analyses reduced the number of possible *APOE*\*4+ gene discoveries. It will thus be important to integrate additional datasets into future *APOE*\*4 stratified GWAS, especially clinically and pathology confirmed case-control cohorts, which will amplify power for cTWAS gene discoveries and improve downstream compound enrichment. Similarly, while the ROSMAP snRNA-seq dataset was large for sc-eQTL analyses, inclusion of other datasets will enable detection of additional heritable genes to be assessed in cTWAS. We also performed AFR and Japanese GWAS, but sample sizes were relatively small and sc-eQTL data for cTWAS were not ancestry matched, such that larger multi-ancestry datasets are needed to better assess cross-ancestry consistency. Regarding the cell-specificity of our findings, it should be noted that while cTWAS implicates likely causal roles for cell-type-gene pairs, the respective genes can still be involved in other cell-types over the disease course. Lastly, while we focused on cell-specific transcriptomics analyses, considering other tissues and gene regulatory features may also be relevant and could expand gene prioritization.

## Conclusions

We identified a set of *APOE*\*4 stratified genes that may be causal for AD through brain cell-type-specific mechanisms, prioritizing top genes for further interrogation. We additionally identified compounds that may be repurposed or shed light on therapeutic avenues for treating AD based on an individual’s *APOE*\*4 status. Top identified compounds such as Hydrocortisone and Trolox implicate oxidative stress and neuroinflammation as potential biological targets in *APOE*\*4+ individuals. Altogether, our study substantially expands current insights into the genetic architecture and molecular etiology of AD, while paving the road towards *APOE*\*4 genotype-tailored therapies.

## Supporting information

eTable

## Data Availability

Data used in the APOE*4 stratified AD GWAS for European cohort and African mixed cohort are available upon application to: dbGaP (https://www.ncbi.nlm.nih.gov/gap/); NIAGADS (https://www.niagads.org/); LONI (https://ida.loni.usc.edu/); AMP-AD knowledge portal / Synapse (https://www.synapse.org/); Rush (https://www.radc.rush.edu/); NACC (https://naccdata.org/); UKB (https://www.ukbiobank.ac.uk/)
The specific data repository and identifiers for ADGC and ADSP data are indicated in eTable1-2 of the supplement. Full APOE*4 GWAS summary statistics per ancestry will be available in NIAGADS and GWAS Catalogue upon publication.
TWAS gene weights and pseudobulk expression count matrices and were generated as previously reported in Fujita et al., Nat Genet 2024 (https://doi.org/10.1038/s41588-024-01685-y). The underlying snRNA-seq data (raw UMI count matrices and cell type annotations) are available from Synapse (https://doi.org/10.7303/syn52335732) upon application. Cell-type-specific eQTL summary statistics used in this study are available from Synapse (https://doi.org/10.7303/syn52335732). The scATAC-seq peak files used in this study were derived from Corces et al., Nat Genet 2020 (https://doi.org/10.1038/s41588-020-00721-x). The raw data are available in the Gene Expression Omnibus under accession number GSE147672. The GMT file for drug enrichment analysis was obtained from DSigDB (https://dsigdb.tanlab.org/).

## Data Availability

Data used in the *APOE*\*4 stratified AD GWAS for European cohort and African mixed cohort are available upon application to:

– dbGaP (https://www.ncbi.nlm.nih.gov/gap/)
– NIAGADS (https://www.niagads.org/)
– LONI (https://ida.loni.usc.edu/)
– AMP-AD knowledge portal / Synapse (https://www.synapse.org/)
– Rush (https://www.radc.rush.edu/)
– NACC (https://naccdata.org/)
– UKB (https://www.ukbiobank.ac.uk/)

The specific data repository and identifiers for ADGC and ADSP data are indicated in **eTable1-2** of the supplement. Full APOE*4 GWAS summary statistics per ancestry will be available in NIAGADS and GWAS Catalogue upon publication.

TWAS gene weights and pseudobulk expression count matrices and were generated as previously reported in Fujita et al., *Nat Genet* 2024 (https://doi.org/10.1038/s41588-024-01685-y). The underlying snRNA-seq data (raw UMI count matrices and cell type annotations) are available from Synapse (https://doi.org/10.7303/syn52335732) upon application. Cell-type–specific eQTL summary statistics used in this study are available from Synapse (https://doi.org/10.7303/syn52335732). The scATAC-seq peak files used in this study were derived from Corces et al., Nat Genet 2020 (https://doi.org/10.1038/s41588-020-00721-x). The raw data are available in the Gene Expression Omnibus under accession number GSE147672. The GMT file for drug enrichment analysis was obtained from DSigDB (https://dsigdb.tanlab.org/).

## Authors’ contributions

Y.Z. and M.E.B. had full access to all the data in the study and take responsibility for the integrity of the data and the accuracy of the data analysis. Y.Z. performed data analyses and interpretation, wrote the manuscript, and contributed to study design. N.C., C.Y., and S.K.S. contributed to data processing and analyses. M.F., Z.A.G., Y.L.G., D.S., K.O., T.M., N.H., A.M., T.I., C.C., V.N., M.R.C., V.M., and M.D.G. contributed data and resources and were involved in discussion of results. C.P., M.R.C, and V.M. contributed to study design and data interpretation. M.E.B. performed data acquisition and analyses, designed analyses, designed study, supervised analyses, supervised work, wrote paper, and obtained funding. All authors contributed to critical revision of the manuscript.

## Role of Funder/Sponsor

The funding organizations and sponsors had no role in the design and conduct of the study; collection, management, analysis, and interpretation of the data; preparation, review, or approval of the manuscript; and decision to submit the manuscript for publication.

## Competing interests

C.C. has received research support from GSK and EISAI. C.C. is a member of the scientific advisory board of Circular Genomics and owns stocks. C.C. is a member of the scientific advisory board of ADmit.

## Acknowledgements

We thank all study participants and their families as well as many involved institutions and their staff. This work was supported by grants from the National Institutes of Aging (R00AG075238, M.E.B) and from the Japan Foundation for Aging and Health (AMED; grant number JP25dk0207060, T.I.).

Data for this study were prepared, archived, and distributed by the National Institute on Aging Alzheimer’s Disease Data Storage Site (NIAGADS) at the University of Pennsylvania (U24-AG041689), funded by the National Institute on Aging. The contents of this article do not represent the views of the National Institutes of Health, the U.S. Department of Veterans Affairs, or the United States Government.

Cohorts specific acknowledgments are provided in the **supplement**.

## Notes

### Author Declarations

The study protocol was granted an exemption by the Washington University Institutional Review Board because the analyses were carried out on de-identified, off-the-shelf data; therefore, additional informed consent was not required.

